# Association of Acculturation and Cardiometabolic Disease Incidence Among Immigrants of South Asian Ancestry in a National Biobank

**DOI:** 10.1101/2022.10.23.22281197

**Authors:** Nikhil Kathiresan, Amit V. Khera, Aniruddh P. Patel

**Affiliations:** Cardiovascular Disease Initiative, Broad Institute of MIT and Harvard, Cambridge, Massachusetts; Milton Academy, Milton, Massachusetts; Verve Therapeutics, Cambridge, Massachusetts; Division of Cardiology, Department of Medicine, Massachusetts General Hospital, Boston, Massachusetts; Center for Genomic Medicine, Department of Medicine, Massachusetts General Hospital, Boston, Massachusetts; Department of Medicine, Harvard Medical School, Boston, Massachusetts; Cardiovascular Research Center, Massachusetts General Hospital, Boston, Massachusetts

## Abstract

**Background:** Individuals of South Asian ancestry experience higher risk of cardiometabolic disease compared with most other groups, and the impact of acculturation on this risk warrants further study.

**Methods:** We studied individuals of South Asian (n=8,420) and European (n=441,696) ancestry free of baseline cardiometabolic disease in the UK Biobank prospective cohort. South Asians were divided into 3 acculturation groups based on time lived in UK: born in UK (n=801); born abroad, in UK >5yrs (n=7213), and born abroad, in UK ≤5yrs (n=406). We estimated the risk for incident coronary artery disease (CAD), type 2 diabetes mellitus, chronic kidney disease, ischemic stroke, and non-alcoholic fatty liver disease, comparing South Asian acculturation groups vs. European ancestry individuals using Cox proportional hazards regression.

**Results:** Over a median follow-up of 12 yrs, individuals of South Asian ancestry, regardless of acculturation group, had a higher risk of cardiometabolic disease when compared with European ancestry individuals. After adjusting for age and sex, the HR for CAD was 1.76 (95%CI 1.26-2.45;p=0.001) for UK-born South Asians; 2.00 (95%CI 1.86-2.16;p<0.001) for South Asians born abroad, in UK >5 yrs; and 2.03 (95%CI 1.42-2.91;p<0.001) for South Asians born abroad, lived in UK ≤5 yrs. Similarly elevated risk across groups was observed for DM and CKD.

**Conclusion:** In a large prospective study, we show that individuals of South Asian (vs. European) ancestry are at higher risk for cardiometabolic disease, regardless of time lived in UK.

## Main Text

Cardiometabolic disease is a leading cause of death globally, and clinical practice guidelines recognize that individuals of South Asian ancestry–who represent a quarter of the world population and comprise fast-growing immigrant communities internationally–are at particularly high risk.^1,2^ It is uncertain whether any observed higher risk varies according to time of immigration.^3^

To address this, we analyzed a prospective cohort study of 8,420 individuals of South Asian ancestry and 441,696 individuals of European ancestry free of baseline cardiometabolic disease in the UK Biobank recruited between 2006-2009 with median follow-up of 12 years.^4^ Individuals of South Asian ancestry were further divided into 3 acculturation groups based on time lived in UK: born in UK (n=801); born abroad, lived in UK >5yrs (n=7213), and born abroad, lived in UK ≤5yrs (n=406). The primary end point was cardiometabolic disease, defined on the basis of hospitalization and death registry records indicating diagnosis codes for coronary artery disease (CAD), type 2 diabetes mellitus (DM), chronic kidney diseases (CKD), ischemic stroke, or nonalcoholic fatty liver disease (NAFLD). Risk for development of incident cardiometabolic disease after enrollment associated with a given acculturation group relative to UK-born European ancestry individuals was computed with Cox proportional hazards regression models including the covariates of age and sex. This research was conducted with the UK Biobank resource under Application No. 7089 and approved by the Mass General Brigham institutional review board.

Among 450,116 included individuals, the mean (SD) age was 57.4 (8.0) and 54.0 (8.5) years for individuals of European and South Asian ancestry, respectively and 54.0% vs. 45.6% percent were female, respectively. At the time of enrollment, baseline characteristics in European vs. South Asian ancestry individuals included: 45,745 (10.4%) vs. 735 (8.8%) current smokers, mean (SD) glycated hemoglobin of 36.0 (6.5) vs. 40.8 (10.5) mmol/mol, mean (SD) body mass index of 27.4 (4.8) vs. 27.2 (4.4) kg/m^2^, mean (SD) systolic blood pressure of 140.1 (19.7) vs. 137.0 (19.6) mmHg, mean (SD) diastolic blood pressure of 82.2 (10.7) vs. 82.5 (10.7) mmHg, mean (SD) low-density lipoprotein cholesterol concentration of 137.8 (33.6) vs. 128.8 (32.8) mg/dL, mean (SD) high-density lipoprotein cholesterol concentration of 56.0 (14.8) vs. 48.6 (12.4) mg/dL, mean (SD) triglyceride concentration of 155.6 (90.9) vs. 174.1 (103.1) mg/dL, mean (SD) sleep time of 7.2 (1.1) vs. 7.1 (1.2) hours, mean (SD) sedentary time of 5.0 (3.8) vs. 5.4 (4.6) hours, and 188,890 (44.5%) vs. 4,066 (56.8%) with a healthy diet, respectively.

Over a median follow-up of 12 years, individuals of South Asian ancestry, regardless of acculturation group, had a higher risk of cardiometabolic disease when compared with European ancestry individuals. After adjusting for age and sex, the HR for CAD was 1.76 (95%CI 1.26-2.45;p=0.001) and incidence rate (IR) was 7.81 events per 1000 person-years (events/1000-PY) (95%CI 5.61-10.89) for UK-born South Asians; HR 2.00 (95%CI 1.86-2.16;p<0.001) and IR 8.9 events/1000-PY (95%CI 8.28-9.57) for South Asians born abroad, in UK >5 yrs; and HR 2.03 (95%CI 1.42-2.91;p<0.001) and IR 9.04 events/1000-PY (95%CI 6.32-12.94) for South Asians born abroad, lived in UK ≤5 yrs. For DM, a gradient in risk was observed according to time spent in the UK, with UK-born South Asians having the lowest risk relative to UK-born Europeans (HR 3.05, 95% CI 2.32-3.99; IR 11.67 events/1000-PY, 95%CI 8.91-15.28) and South Asians born abroad, lived in UK ≤5 yrs having the highest relative risk (HR 7.01 95% CI 5.57-8.83; IR 26.69 events/1000-PY, 95% CI 21.21-33.59). Similarly elevated risk across South Asian acculturation groups was observed for CKD, however, no significant differences were noted for risk of ischemic stroke or NAFLD.(Figure 1) After adjusting for traditional risk factors, the HR for CAD was 1.67 (95%CI 1.12-2.50;p=0.012) for UK-born South Asians; 2.00 (95%CI 1.83-2.18;p<0.0001) for South Asians born abroad, in UK >5 yrs; and 2.39 (95%CI 1.57-3.63;p<0.0001) for South Asians born abroad, lived in UK ≤5 yrs. Similar trends were seen after adjusting for these variables for the other cardiometabolic traits.

**Figure 1:**
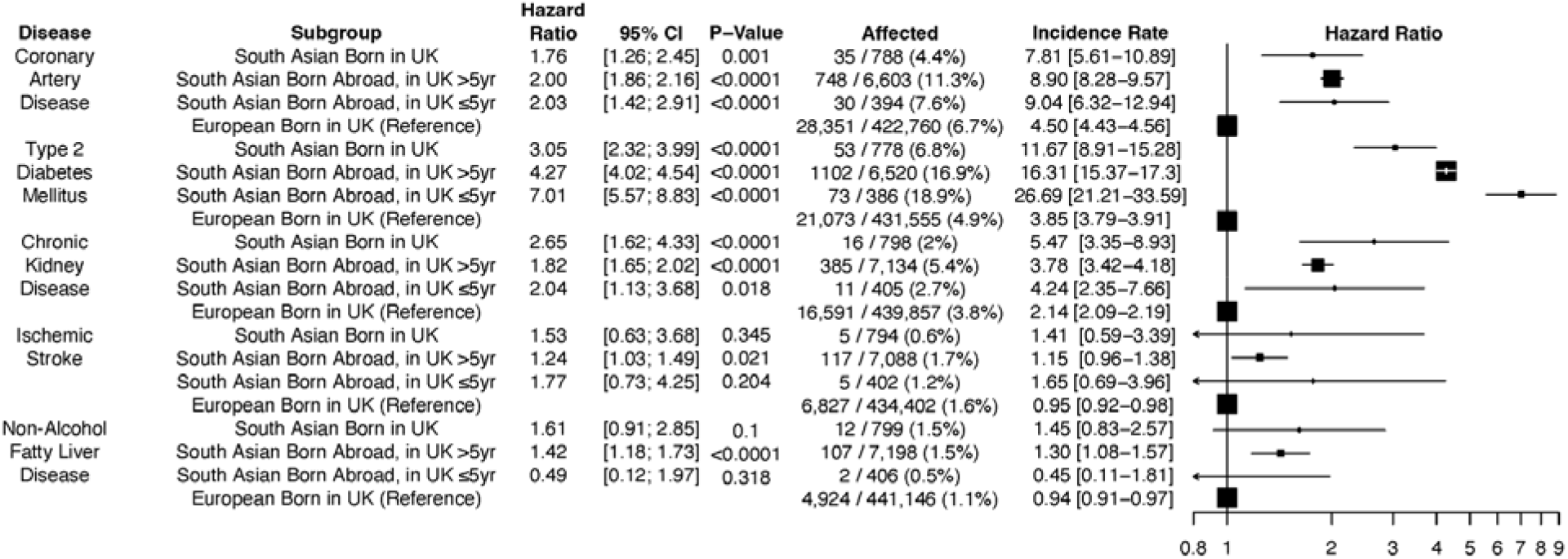
Adjusted hazard ratios of cardiometabolic diseases for South Asian relative to European ancestry individuals stratified by subgroups of time spent in UK

These results build on previous efforts to understand the nature of cardiometabolic risk in South Asian individuals in several key ways. First, the degree of acculturation has limited effect on the higher risk of cardiometabolic diseases outside of diabetes in individuals of South Asian vs. European ancestry. Although the variables determining diabetes risk may differ with more time spent in the UK, these do not translate into increased risk of CAD or CKD. Second, we did not observe a significant heterogeneity in risk factor burden among the three South Asian immigrant groups, suggesting that the effects of acculturation on lifestyle and risk factors variables is also modest. Finally, residual unexplained risk remained after accounting for traditional risk factors, suggesting that other factors common to all three South Asian acculturation groups may be at play. Mitigating this risk may involve targeting other non-traditional factors and further research is warranted into such interventions.^5^

Adjusted hazard ratios with corresponding 95% confidence intervals and p values for coronary artery disease (composite of myocardial infarction, revascularization, and atheroscleroris), type 2 diabetes mellitus, ischemic stroke, chronic kidney disease, and non-alcoholic fatty liver disease, comparing individuals of South Asian ancestry stratfied by time spent in UK to individuals of European ancestry, calculated using Cox proportional hazards regression models with covariates of enrollment age and sex. Adjusted incidence rates estimated as events per 1000 person-years (PY) of follow up time and adjusted for age and sex using Poisson regression.

## Data Availability

All data analyzed in the present study are available upon reasonable request and application to the UK Biobank.

https://biobank.ndph.ox.ac.uk/showcase/

